# Evaluating binary classifiers: extending the Efficiency Index

**DOI:** 10.1101/2022.02.02.22270139

**Authors:** Andrew J Larner

**Author notes:** Correspondence: AJ Larner, Cognitive Function Clinic, Walton Centre for Neurology and Neurosurgery, Lower Lane, Fazakerley, Liverpool, L9 7LJ, United Kingdom.

## Abstract

An “efficiency index” (EI) for the evaluation of binary classifiers was recently characterised, where EI is the ratio of classifier accuracy to inaccuracy. The purpose of this study was to further develop EI by substituting balanced accuracy and unbiased accuracy in place of accuracy, and their respective complements in place of inaccuracy, to construct balanced EI and unbiased EI measures. Additional investigations, using the dataset of a prospective pragmatic test accuracy study of a cognitive screening instrument, explored use of the log method to calculate confidence intervals for the various EI formulations; the dependence of EI formulations on prevalence; and comparison of EI formulations with analogous formulations based on the Identification Index (II), a previously described metric which is also based on accuracy and inaccuracy, where II is accuracy minus inaccuracy. EI formulations are shown to have advantages over II formulations, in particular their boundary values (0 and ∞) mean that negative values never occur, unlike the case for II, and the inflection point of 1 demarcates likelihood of correct versus incorrect classification.

## 1. Introduction

Many metrics exist for the evaluation of binary classifiers, with different measures being favoured in different disciplines. For example, the true positive and true negative rates (TPR, TNR) derived from the standard 2×2 contingency table, also known as Sensitivity and Specificity, are often used in the evaluation of medical screening and diagnostic tests,^1^ whereas the F measure, also known as F or F1 score,^2^ and Matthews’ correlation coefficient (MCC)^3^ are often preferred in data retrieval and machine learning contexts. Unlike the paired measures of TPR and TNR, F measure and MCC are unitary, global measures of test accuracy, other examples of which include Accuracy (Acc) and the area under the receiver operating characteristic curve (AUC ROC).

Recently a further metric, the efficiency index (EI), was proposed as a measure to evaluate binary classifiers.^4^ (Note that this EI differs from the similarly named physical index of speed achieved in relation to power output, and various other efficiency indexes described for energy efficiency or in business and finance.) EI was based on Acc and its complement, Inaccuracy (Inacc), and defined as:

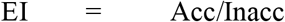

Acc is a weighted average of TPR and TNR, with weights related to sample prevalence (P and 1 – P or P’):

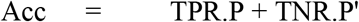

Similarly, Inacc is a weighted average of false positive rate (FPR) and false negative rate (FNR), with weights related to P and P’:

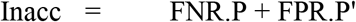

where FNR and FPR are the complements of TPR and TNR respectively. Hence, EI may be rewritten as

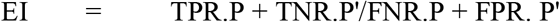

which indicates the dependence of EI on prevalence.

EI has been suggested to have some advantages over other unitary measures of binary classification.^4^ EI has boundary values of 0 and ∞, with an inflection point at 1, where a value >1 indicates correct classification and a value of <1 indicates incorrect classification, such that values >>1 are desirable and a value of ∞ is optimal. These boundary values and inflection point are shared with likelihood ratios (LRs) and it has previously been shown that EI may be characterised using the same qualitative and quantitative systems of classification used for LRs.^5,6^ This may be of advantage when communicating risk, for example of a medical test producing correct diagnosis rather than misdiagnosis.^4^

The purpose of this study was to extend consideration of EI in various ways. Firstly, by using other formulations of Acc, namely Balanced Accuracy and Unbiased Accuracy,^7^ new Balanced EI and Unbiased EI formulations have been developed. Secondly, confidence intervals for these EI formulations have been calculated using the same logarithmic method used for LRs and for diagnostic odds ratios (DORs). Thirdly, the dependence of EI formulations on prevalence has been examined.

Fourthly, the EI metrics have been compared with another, previously described, metric based on Acc and Inacc, the Identification Index (II),^8^ which was also further developed using Balanced Accuracy and Unbiased Accuracy to produce Balanced II and Unbiased II formulations. Worked examples of all these developments of EI are examined using the dataset of a test accuracy study for a dementia screening test.

## 2. Material and methods

The utility of EI was previously shown^4^ using published data from a prospective test accuracy study of a screening test for dementia and cognitive impairment, the Mini-Addenbrooke’s Cognitive Examination (MACE).^9^ The same dataset^10^ was used here, from a study in which subjects gave informed consent and study protocol was approved by the institute’s committee on human research (Walton Centre for Neurology and Neurosurgery Approval: N 310).

### 2.1 Extended formulations of EI

Balanced Accuracy (BAcc) has been defined^11^ as:

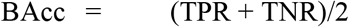

BAcc has been advocated in place of Acc when examining biased classifiers and/or imbalanced datasets, i.e. where P and hence P’ are markedly different from 0.5.^7,11^

Using BAcc in place of Acc, one may characterise a Balanced Efficiency Index (BEI) as:

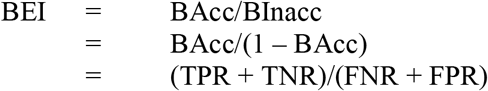

Hence unlike EI, BEI is independent of prevalence.

Unbiased Accuracy (UAcc) has been defined^12^ as:

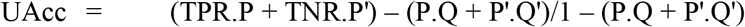

where Q is the level or bias of the test.^13^ This formulation corrects for the proportion of agreement expected by chance alone, equating to Cohen’s kappa statistic, κ.^7,12^

Using UAcc in place of Acc, one may characterise an Unbiased Efficiency Index (UEI) as:

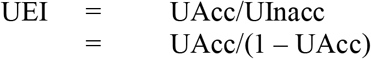

Like EI, BEI and UEI have boundary values of 0 and ∞, with an inflection point at 1, where a value >1 indicates correct classification and a value of <1 indicates incorrect classification, such that values >>1 are desirable and a value of ∞ is an optimal classifier.

### 2.2 Confidence intervals for EI formulations

EI, BEI, and UEI are all of the form a/(1 – a) and hence are odds ratios. (They could all also be expressed as (1 – a)/a, hence as odds against ratios.) It is possible to calculate confidence intervals (CI) for EI, BEI, and UEI, as for LRs and DORs, through a logarithmic transformation.^14^ Applying the log method to data from the four cells of the 2×2 contingency table (where true positive = a, false positive = b, false negative = c, true negative = d), for the 95% CI the formula for EI is:

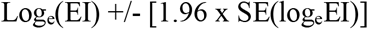

where:

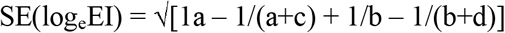

With the appropriate substitutions, this method may also be used to calculate 95% CI for BEI and UEI.

### 2.3 Dependence of EI formulations on prevalence

As BEI is based solely on columnar ratios from the 2×2 contingency table (TPR, FNR, TNR, FPR), it is independent of P. Whilst this might be deemed an advantage, the biasing effects of random associations between classifier result and prevalence may be allowed for by taking into account P, as is the case in EI and UEI. This may be examined by rescaling Acc and UAcc (and hence Inacc and UInacc) across a range of prevalence (P) values at a fixed value of Q (i.e. fixed classifier cut-off, and hence fixed values of TPR, TNR, FNR and FPR) and hence calculating rescaled EI and UEI values from the respective equations.

### 2.4 Comparison of EI formulations with II formulations

An Identification Index (II) has been defined^8^ as:

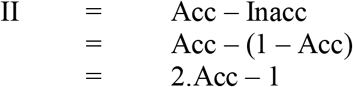

Hence whereas Inacc is the divisor of Acc in EI, it is the subtrahend of Acc in II.

By analogy with EI and BEI, one may characterise a Balanced Identification Index (BII) as:

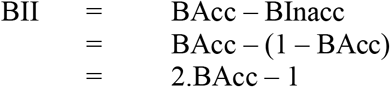

However, this formulation contributes no additional information, since: BAcc = (TPR + TNR)/2

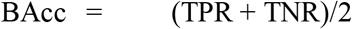

Hence:

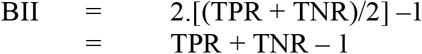

This is the formula for the Youden index or J statistic,^15^ also termed bookmarker informedness,^3^ the value of which is immediately available from TPR and TNR.

By analogy with EI and UEI, one may also characterise an Unbiased Identification Index (UII) as:

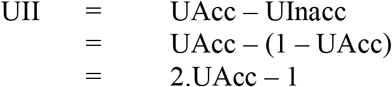

II, BII and UII have boundary values of −1 and +1, unlike the various efficiency indexes. They have an inflection point at 0, values above which imply the classifier is useful rather than useless, with a value of 0 denoting a random classifier, a value of 1 denoting an optimal classifier, and values <0 indicating that the classifier is misleading.

## 3. Results

### 3.1 Extended formulations of EI

Calculations of EI, BEI, and UEI are shown in Table 1 and illustrated in Figure 1. These parameters showed different maxima (EI at MACE cut-off ≤14/30; BEI ≤20/30; UEI ≤15/30). The plots of EI and BEI cross (EI = BEI somewhere between the cut-offs of ≤18/30 and ≤19/30 at this prevalence) but neither cross the plot of UEI which lies beneath them. So stringent is UEI that it has a value >1 only at its maximum (≤15/30).

**Table 1:** Diagnosis of dementia: comparing EI, BEI, and UEI metrics at various MACE cut-offs (fixed value of P = 0.151)

| **Cut-off** | **EI** | **BEI** | **UEI** |
| --- | --- | --- | --- |
| ≤29/30 | 0.204 | 1.045 | 0.006 |
| ≤28/30 | 0.246 | 1.101 | 0.015 |
| ≤27/30 | 0.355 | 1.283 | 0.043 |
| ≤26/30 | 0.507 | 1.538 | 0.081 |
| ≤25/30 | 0.716 | 1.882 | 0.135 |
| ≤24/30 | 0.982 | 2.289 | 0.198 |
| ≤23/30 | 1.274 | 2.759 | 0.272 |
| ≤22/30 | 1.668 | 3.310 | 0.368 |
| ≤21/30 | 2.199 | 3.854 | 0.484 |
| ≤20/30 | 2.813 | 4.236 | 0.605 |
| ≤19/30 | 3.364 | 4.181 | 0.689 |
| ≤18/30 | 4.033 | 4.000 | 0.776 |
| ≤17/30 | 4.207 | 3.525 | 0.745 |
| ≤16/30 | 5.292 | 3.785 | 0.934 |
| ≤15/30 | 6.123 | 3.484 | 1.012 |
| ≤14/30 | 6.550 | 2.922 | 0.980 |
| ≤13/30 | 6.475 | 2.425 | 0.795 |
| ≤12/30 | 6.475 | 2.195 | 0.718 |
| ≤11/30 | 6.260 | 1.874 | 0.567 |

Abbreviations: EI = efficiency index, BEI = balanced efficiency index, UEI = unbiased efficiency index

**Figure 1:**
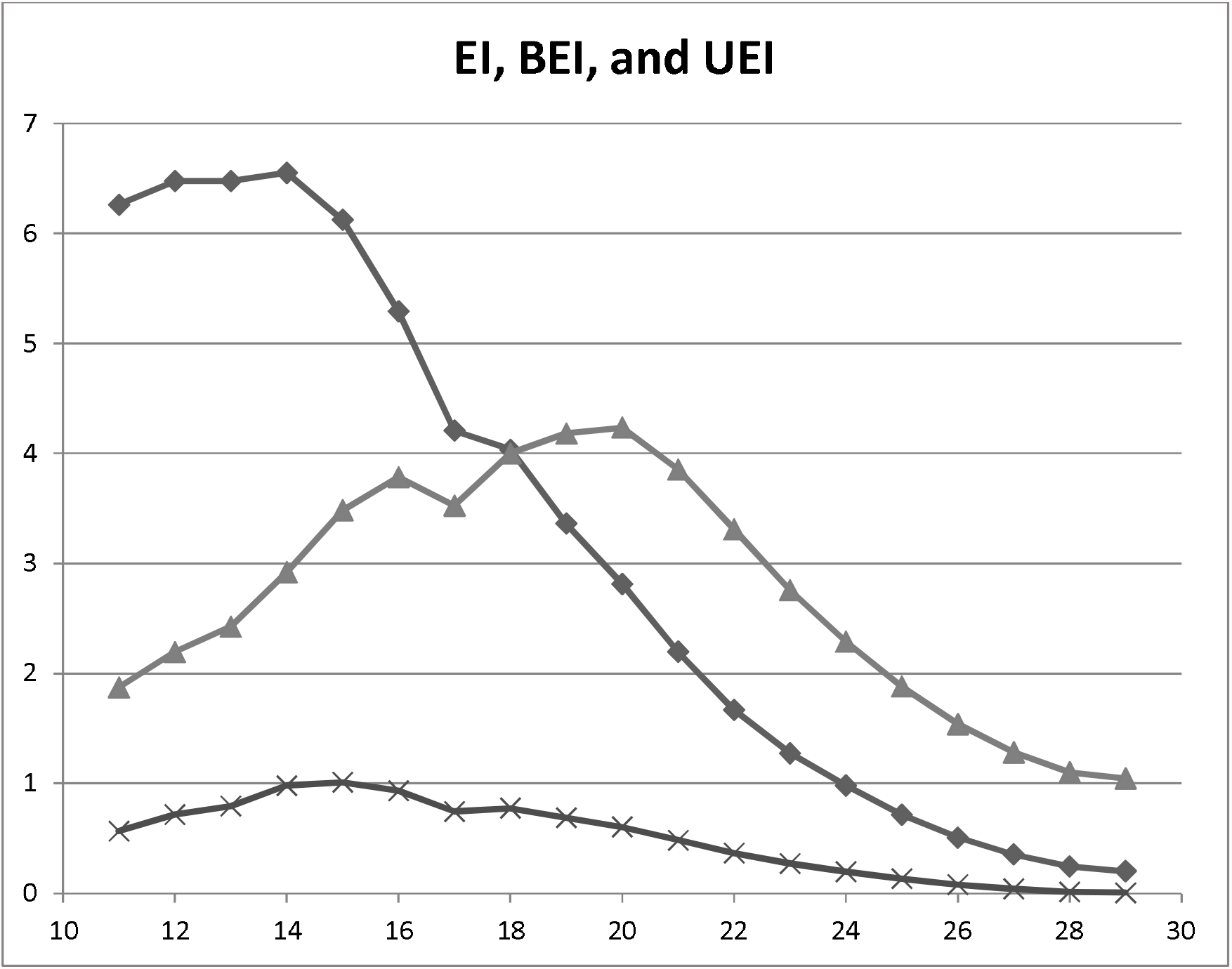
Plot of EI (♦), BEI (▴) and UEI (×) values (y axis) vs MACE cut-off score (x-axis) Abbreviations: EI = efficiency index, BEI = balanced efficiency index, UEI = unbiased efficiency index

### 3.2 Confidence intervals for EI formulations

The 95% confidence intervals (95% CI) for EI, BEI, and UEI at the previously defined optimal MACE cut-off in the test dataset (≤20/30)^10^ were calculated by the log method as follows:

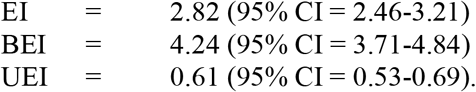

At their respective maxima, the values for EI and UEI were 6.55 (95% CI = 4.85-8.84) and 1.01 (95% CI = 0.78-1.32). Thus, even at its optimal cut-off, the 95% CI for UEI crossed 1.

### 3.3 Dependence of EI formulations on prevalence

The dependence of EI, BEI, and UEI on prevalence are shown in Table 2 and illustrated in Figure 2. By definition BEI is independent of P, hence does not vary, whereas EI and UEI gradually increase with increasing prevalence, since when P = 1 then EI and UEI = ∞ because the denominators in their respective defining equations = 0, and when P = 0 then EI and UEI = 0 because the numerators in their respective defining equations = 0.

**Table 2:**
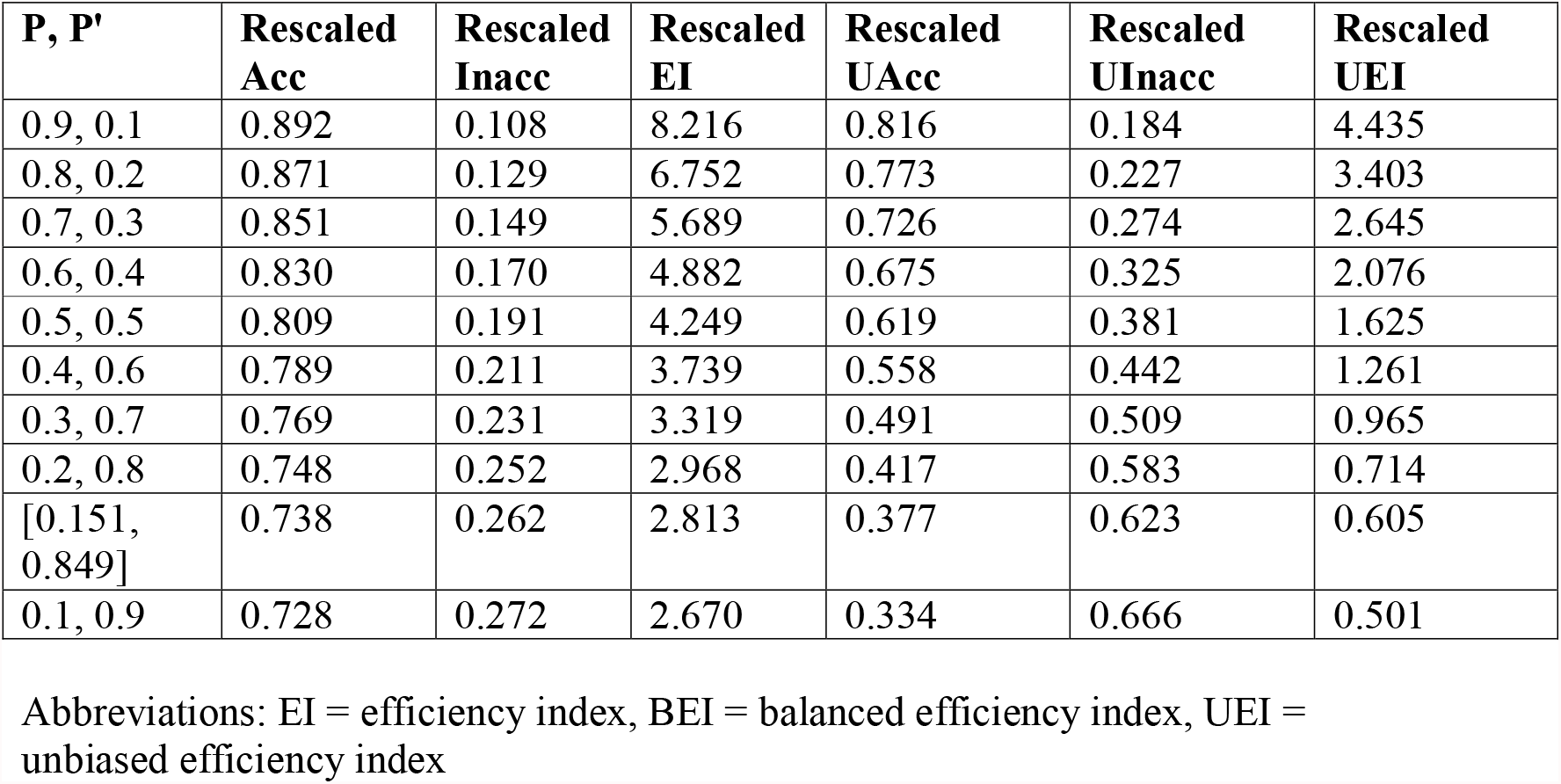
Rescaled values of EI and UEI at varying values of P for diagnosis of dementia using MACE at fixed cut-off (≤20/30; Q = 0.387, Q’ = 0.613) and hence Sens (0.912) and Spec (0.707), and hence fixed BEI (= 4.236)

**Figure 2:**
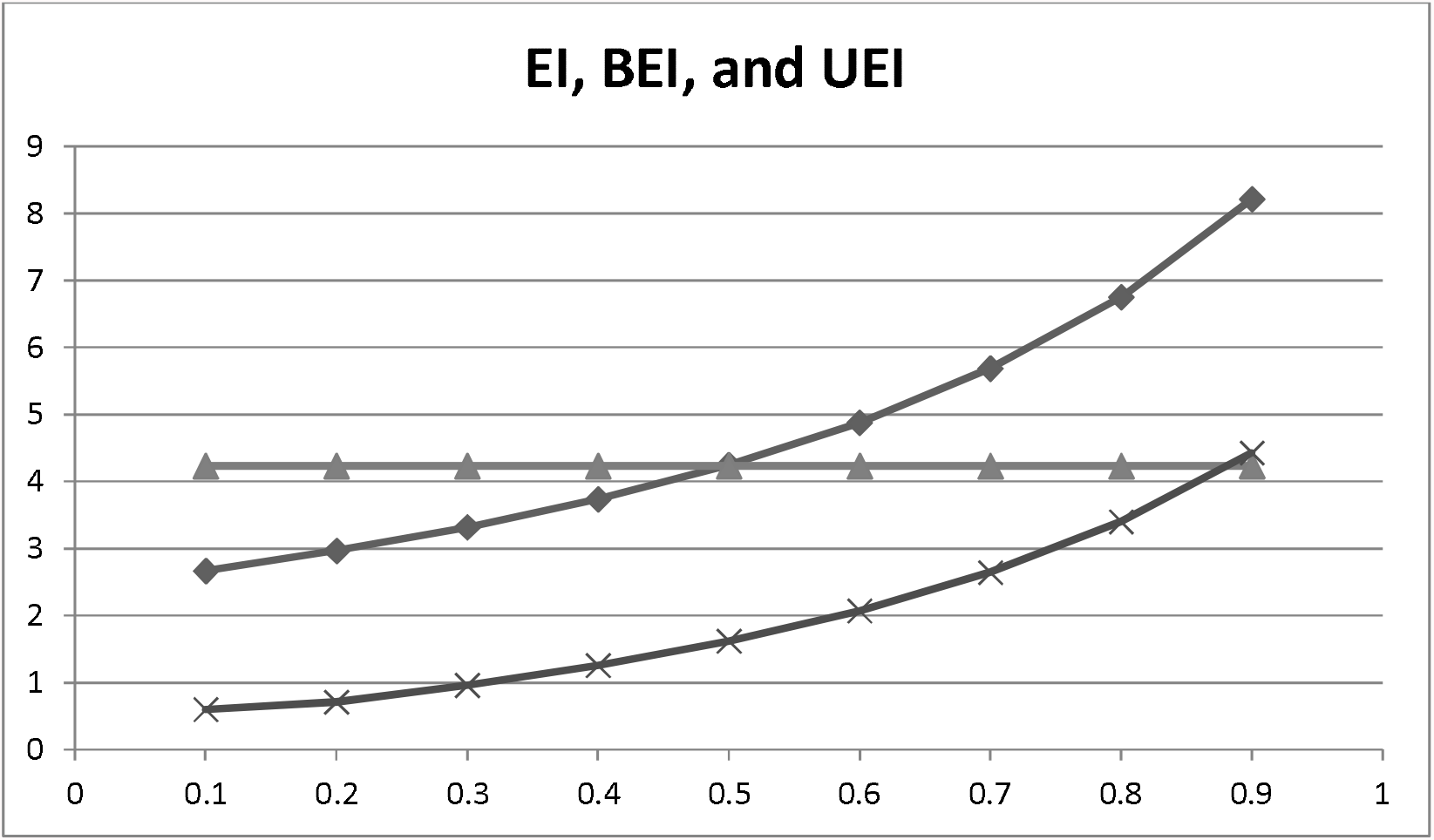
Plot of rescaled EI (♦) and UEI (×) values with fixed BEI (▴) (y axis) vs prevalence (x-axis)

### 3.4 Comparison of EI formulations with II formulations

Calculations of II, BII, and UII are shown in Table 3 and illustrated in Figure 3 (note the different y-axis scale from Figure 1). These parameters showed different maxima (II ≤14/30; BII ≤20/30; UII ≤15/30) which corresponded to the maxima for the corresponding EI formulations. The plots of II and BII cross (II = BII = Y somewhere between the cut-offs of ≤18/30 and ≤19/30 at this prevalence) but neither cross the plot of UII which lies beneath them, as was observed for the various EI formulations. So stringent is UII that it has a value >0 only at its maximum (≤15/30).

**Table 3:** Diagnosis of dementia: comparing II, BII, and UII metrics at various MACE cut-offs (fixed value of P = 0.151)

| **Cut-off** | **II** | **BII** | **UII** |
| --- | --- | --- | --- |
| ≤29/30 | -0.660 | 0.022 | -0.988 |
| ≤28/30 | -0.606 | 0.048 | -0.970 |
| ≤27/30 | -0.476 | 0.124 | -0.918 |
| ≤26/30 | -0.328 | 0.212 | -0.850 |
| ≤25/30 | -0.166 | 0.306 | -0.762 |
| ≤24/30 | -0.010 | 0.392 | -0.670 |
| ≤23/30 | 0.120 | 0.468 | -0.572 |
| ≤22/30 | 0.250 | 0.536 | -0.462 |
| ≤21/30 | 0.374 | 0.588 | -0.348 |
| ≤20/30 | 0.476 | 0.618 | -0.246 |
| ≤19/30 | 0.542 | 0.614 | -0.184 |
| ≤18/30 | 0.608 | 0.600 | -0.126 |
| ≤17/30 | 0.616 | 0.558 | -0.146 |
| ≤16/30 | 0.682 | 0.582 | -0.034 |
| ≤15/30 | 0.718 | 0.554 | 0.006 |
| ≤14/30 | 0.734 | 0.490 | -0.010 |
| ≤13/30 | 0.732 | 0.416 | -0.114 |
| ≤12/30 | 0.732 | 0.374 | -0.164 |
| ≤11/30 | 0.724 | 0.304 | -0.276 |

Abbreviations: II = identification index, BII = balanced identification index, UII = unbiased identification index

**Figure 3:**
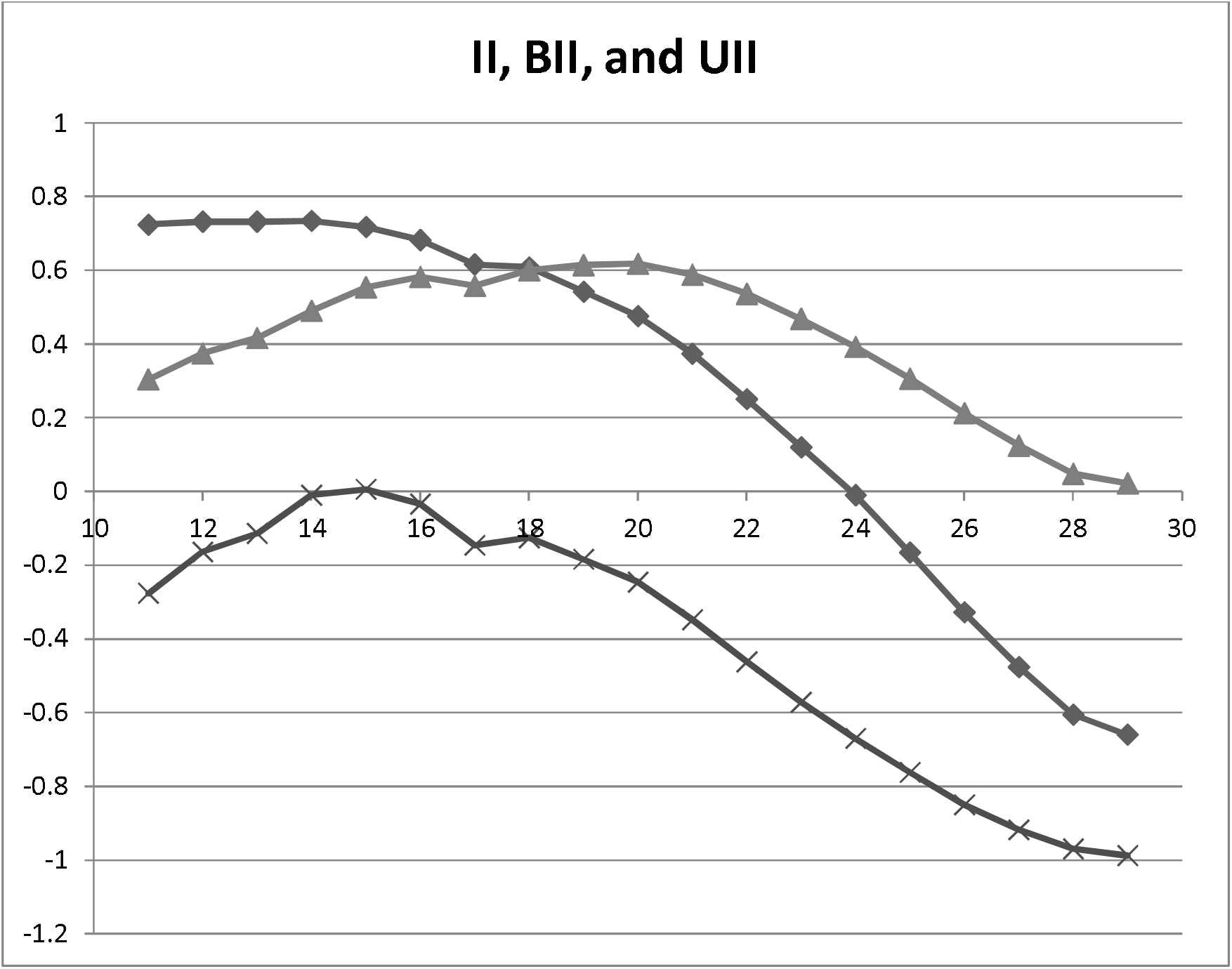
Plot of II (♦), BII (▴) and UII (×) values (y axis) vs MACE cut-off score (x-axis)

## 4. Discussion

This study aimed to develop the recently proposed efficiency index, EI, as a metric for the evaluation of binary classifiers by application of different formulations of accuracy, namely balanced accuracy and unbiased accuracy. This has extended the formulation of EI to BEI and UEI by using BAcc and UAcc in place of Acc respectively. By the same procedure, the formulation of the identification index, II, has been extended to BII and UII.

Of the EI formulations, BEI was found to be a relatively more liberal or optimistic metric than EI, whereas UEI was a relatively more conservative or pessimistic metric, as was previously shown to be the case for UAcc relative to Acc.^7^ In the calculated example, only one UEI point had a value >1, and this may be a more generalised problem because of the conservative values of UAcc. This arises because UEI, like UAcc, takes into account values of both P and Q, hence removing the biasing effects of random associations between, in this particular case, the test result and disease prevalence.

Confidence intervals for EI formulations were easily calculated from the 2×2 contingency table base data using the logarithmic method.^14^ In the worked examples, the confidence intervals for EI and BEI at the optimal MACE cut-off were greater than 1, suggesting that the test under examination was more likely to classify correctly than incorrectly, whereas for UEI even at its maximal cut-off the confidence intervals crossed 1, suggesting that when using this stringent metric this classifier is no more likely to classify correctly than to misclassify.

By definition BEI, like BAcc, is an unscaled measure, calculation of which is independent of P. EI and UEI formulations vary with P, by definition. This is because EI and UEI, like Acc and UAcc, take into account the value of P, hence removing the biasing effects of random associations between test result and disease prevalence. EI occupies a middle ground between BEI and UEI, since it takes into account P but not Q.^4^ In the worked examples, both EI and UEI increased with P for the classifier under examination. Using the same dataset, a previously described “likelihood to be diagnosed or misdiagnosed” (LDM), which has some overlap with EI,^4^ showed a fall in value at either very low or very high disease prevalence using the same cut-off.^16^

Of the II formulations, BII simplifies to an existing metric, the Youden index, Y, or bookmarker informedness, and hence contributes no new information. As it is based on strictly columnar ratios from the 2×2 contingency table (TPR, TNR), it is, at least algebraically, independent of P, although in practice TPR and TNR may be found to vary according to the heterogeneity of the sampled population.^17^ As UII is based on UAcc, it is by definition dependent on P and Q, with II occupying a middle ground (taking into account P but not Q).

EI formulations have advantages of over II formulations. The range of the EI formulations (boundary values 0 and ∞) means that negative values never occur, as is also the case for LRs and DORs. EI values are easily classifiable, either qualitatively or quantitatively,^4^ using existing classification schemes for LRs,^5,6^ with an inflection point at 1, denoting a random classifier, above which correct classification is favoured over incorrect classification. The qualitative classification scheme for odds ratios proposed by Rosenthal might also be applicable to EI formulations (small □1.5, medium □2.5, large □4, and very large □10 or greater).^18^

In contrast to EI formulations, the range of the II formulations (boundary values −1 and +1) means that negative values can occur (see Table 3 and Figure 3), the meaning of which may be unclear. There is no evident classificatory scheme for these metrics, other than larger better, with an inflection point at 0, values above which imply the result is useful rather than useless, and below which imply the result is misleading. In the worked example, only one UII point had a value >0, and this may be a more generalised problem because of the conservative values of UAcc. These shortcomings may account, at least in part, for the fact that the original II formulation has been rarely adopted or examined.^1,15,19^

It has previously been suggested that EI may have greater utility than other metrics in communicating risk of test outcomes^4^ and this may also be true of the extended EI formulations developed here, although this requires empirical examination. Other future studies might also examine the relationship between EI formulations as possible indexes of effect size which might be qualitatively related to Cohen’s d.^20^

In summary, EI, BEI and UEI appear to be serviceable metrics for the evaluation of binary classifiers. They are easily calculated from the cells of the 2×2 contingency table, as are their confidence intervals using the log method. They are easily classified qualitatively and quantitatively. Of the various extensions of EI examined here, UEI is the most stringent in that it corrects for prevalence and threshold and therefore may prove to be the most appropriate of these measures to be used in the assessment and comparison of the performance of different classifiers.

## Data Availability

All data produced in the present work are contained in the manuscript

## Funding and Acknowledgements

None

